# Added-value of whole exome and RNA Sequencing in advanced and refractory cancer patients with no molecular-based treatment recommendation based on a 90-gene panel

**DOI:** 10.1101/2022.02.08.22270301

**Authors:** Armelle Dufresne, Valéry Attignon, Anthony Ferrari, Laurie Tonon, Séverine Tabone-Eglinger, Philippe Cassier, Olivier Trédan, Nadège Corradini, Armelle Vinceneux, Aurélie Swalduz, Alain Viari, Sylvie Chabaud, David Pérol, Jean Yves Blay, Pierre Saintigny

## Abstract

**Importance:** While comprehensive tumor molecular profile by whole exome and RNA sequencing (WES/RNA-Seq) is now feasible in routine practice, it remains unclear whether this increases therapeutic options as compared to a more limited targeted gene panel (TGP) plus array-based comparative genomic hybridization (aCGH) in advanced cancer patients.

**Objective:** To determine the added value of WES/RNA-Seq in advanced and refractory cancer patients who had no molecular-based treatment recommendation (MBTR) based on a TGP/aCGH in the course of a clinical trial.

**Design:** Retrospective analysis.

**Setting:** Single center.

**Participants:** We selected 50 patients previously included in the PROFILER trial (NCT01774409) for which no molecular-based therapy could be recommended in the course of the clinical trial based on a targeted 90-gene panel and aCGH. For each patient, the frozen tumor sample mirroring the FFPE sample used for TGP/aCGH analysis were processed for WES and RNA-Seq. Data from TGP/aCGH were reanalyzed and together with WES/RNA-Seq, findings were simultaneously discussed at a new molecular tumor board (MTB).

**Main outcomes and Measures:** MBTR based on TGP/aCGH versus WES/RNA-Seq were compared.

**Results:** After exclusion of variants of unknown significance, a total of 167 somatic molecular alterations were identified in 50 patients (median: 3; range: 1-10). Out of these 167 relevant molecular alterations reported by the biologist, 51 (31%) were common to both TGP/aCGH and WES/RNA-Seq, 19 (11%) were identified by the TGP/aCGH only and 97 (58%) were identified by WES/RNA-Seq only, including 2 fusion transcripts in two patients. A MBRT was provided in 4/50 (8%) patients using the information from TGP/aCGH vs. 9/50 (18%) patients using WES/RNA-Seq findings. Three patients had similar recommendations based on TGP/aCGH and WES/RNA-Seq.

**Conclusion and Relevance:** In advanced and refractory cancer patients in whom no MBRT was recommended from TGP/aCGH, WES/RNA-Seq allowed to identify more alterations which may in turn, in a limited fraction of patients, lead to new MBRT.

**Key Points:** *Question:* Does WES/RNA-Seq provide additional targeted treatment guidance for advanced cancer patients with no molecular-based treatment recommendation (MBTR) from a 90-tumor gene panel (TGP) sequencing and array-based comparative genomic hybridization (aCGH)?

*Findings:* For fifty advanced cancer patients included in the PROFILER trial with no treatment recommendation based on a TGP/aCGH, frozen tumor sample was processed for WES and RNA-Seq. MBTR was given in 4/50 (8%) patients using the reanalyzed TGP/aCGH vs. 9/50 (18%) patients using WES/RNA-Seq findings.

*Meanings:* WES/RNA-Seq increased the number of patients with MBTR as compared to a TGP/aCGH screening to yet only a minority of patients.

## Introduction

The concept of tumor-agnostic precision oncology is now integrated in routine for a limited set of somatic molecular alterations. [1-3]. In studies assessing the throughput of tumor molecular analysis for patients with advanced solid tumor, the proportion of patients treated with molecular-based therapy ranged from 6% to 26% [4-7]. This low proportion may have prevented these trials to conclude on the benefit of agnostic precision oncology [8-10]. In contrast, several meta-analysis studies reported a significant benefit of a genomic-driven personalized approach to drive patients in phase I and II trials. Extending the molecular analysis to the entire exome may increase the proportion of actionable molecular alterations, of molecular-based treatment recommendations (MBTR) and eventually, of treated patients.

To determine to which extent a whole exome and RNA sequencing (WES/RNA-Seq) analysis increases the proportion of patients with MBTR, a retrospective analysis was conducted in a subset of 50 patients who had available germ line DNA and fresh frozen tumor mirroring the FFPE sample used for Tumor gene Panel and array-based Comparative Genomic Hybridization (TGP/aCGH) analysis and had no MBTR based on the TGP/aCGH [7].

## Methods

### Patients, sample qualification and molecular analysis

The study was conducted at Centre Léon Bérard, was approved on 2/2/2018 by the institutional review board, and was conducted in compliance with the Declaration of Helsinki and Good Clinical Practice guidelines.

We retrospectively selected 50 patients among the 2,579 patients included in the previously reported PROFILER molecular screening program (NCT01774409) who had no MBTR based on the TGP/aCGH during the course of the trial and for whom a fresh frozen tumor mirroring the FFPE sample together with germ line DNA were available in Centre Léon Bérard certified Biobank (BB-0033-00050) [7]. Fresh frozen surgically resected tumor specimens mirroring the FFPE sample were evaluated by an experienced pathologist for tumor cell content ≥ 30% was required. The first 50 cases achieving those criteria were included in the study.

The molecular analysis conducted in PROFILER trial was reported elsewhere [7]. Details on WES/RNA-Seq sequencing and bioinformatics analysis is provided in Supplementary Methods.

### Variants interpretation and treatment recommendation

Analysis pipelines are regularly updated overtime, TGP raw data for the 50 selected patients were thus reanalyzed and a new report issued. Both the TGP and WES/RNA-Seq reports were presented at the Molecular Tumor Board (MTB). The interpretation of somatic single nucleotide variants (SNV) was focused on their clinical impacts and categorized into five TIERs according to the ESMO Scale for Clinical Actionability of molecular Targets (ESCAT) classification [11] (Supplementary Figure 1). MTB presentation was done at the same time to ensure similar treatment options for both tests.

### Statistical analysis

Statistical analyses were conducted with SPSS 23.0 package (IBM, Paris, France). The proportion of variants in each Tier of the ESCAT classification identified with TGP/aCGH versus WES/RNA-Seq was compared using a Fisher’s exact test. A P value of .05 was considered significant.

## Results

The cohort of 50 patients included 14 different histological subtypes of cancer (Table 1). They were comparable to the overall population of the PROFILER study.

**Table 1.** Clinical and pathological characteristics of the patients included in the study. Abbreviations: MBRT: molecular-based recommended therapies.

|  | **Subset of patients included in the study**<br>**(N=50)** |
| --- | --- |
| **Age at diagnosis**<br>**Median (range)** | 54.0 (21.0-81.0) |
| **Gender**<br>**Male**<br>**Female** | 22 (44%)<br>28 (66%) |
| **ECOG-PS**<br>**Missing data**<br>**0**<br>**1**<br>**≥ 2** | 5 (10%)<br>12 (24%)<br>31 (%)<br>2 (4%) |
| **Delay from the date of diagnosis of**<br>**noncurable disease to inclusion (years)**<br>**Median (range)** | 1.0 (0-11.4) |
| **Primary tumor site**<br>**Breast**<br>**Ovary**<br>**Colorectal**<br>**Sarcoma**<br>**Head & Neck**<br>**Other** | 9 (18%)<br>8 (16%)<br>6 (12%)<br>6 (12%)<br>5 (10%)<br>16 (32%) |

After exclusion of 4 (TGP) and 9,619 (WES) variants of unknown significance, TGP and WES identified 52 SNVs and 121 indels. Respectively 70 and 148 molecular alterations including SNVs (n=135, 80%), CNVs (n=29, 17%), one indel (n=1, <1%), one tumor mutational burden (TMB) > 10 mutations per megabase (median TMB: 1, range: 0-24.5) and fusion transcripts (n=2, 1-2%) were reported by the biologist with TGP/aCGH (median per patient 1, range 0-6) and WES/RNA-Seq (median per patient 2, range 0-8). Out of 167 molecular alterations, 51 (30%) were common to both TGP/aCGH and WES/RNA-Seq, 19 (11%) were identified by the TGP/aCGH only and 97 (58%) were identified by WES/RNA-Seq only. Among the latest, two patients were found with a fusion gene by RNAseq (COL1A1::PDGFB or PAX5::FOXP1) that were already known from the initial diagnostic workup. More ESCAT TIER IV and X molecular alterations were identified by WES/RNA-Seq (Table 2).

**Table 2.** Frequency of molecular alterations identified with the 90-gene TGP/aCGH or WES/RNA-Seq the classified according to ESCAT [11]. Proportion of variants in each Tier of the ESCAT classification identified with TGP/aCGH versus WES/RNA-Seq was compared using a Fisher’s exact test (*P* = 0.0154).

|  | **90-gene TGP/aCGH** | **WES/RNA-Seq** |
| --- | --- | --- |
| **TIER I** | 3 (4%) | 6 (4%) |
| **TIER II** | 13 (19%) | 17 (12%) |
| **TIER III** | 33 (47%) | 44 (30%) |
| **TIER IV** | 10 (14%) | 33 (22%) |
| **TIER X** | 11 (16%) | 48 (32%) |
| **Total** | **70 (100%)** | **148 (100%)** |

Whether MBTR differed when they were based on TGP/aCGH vs WES/RNA-Seq was discussed at the MTB (Figure 1). A MBTR was recommended in 4/50 (8%) patients using the information from TGP/aCGH vs. 9/50 (18%) patients using WES/RNA-Seq findings. Three patients had similar recommendations (PI3K/Akt/mTOR inhibitor and KRAS G12C inhibitor in two and one cases respectively) based on either TGP/aCGH or WES/RNA-Seq (Figure 1). The six MBTR exclusively provided by WES/RNA-Seq were 1) a PKC inhibitor for a choroidal melanoma with a GNAQ SNV (not included in the TGP panel, 2) a KIT inhibitor for a gastrointestinal stromal tumor with a KIT D820E mutation (region not covered by TGP), 3) an immune therapy based on a high TMB on WES (not available on TGP) for a malignancy of unknown origin, 4) a PARP inhibitor based on a BRCA loss not identified by TGP for a serous ovarian cancer and 5) and 6) were recommended a PI3K/Akt/mTOR inhibitor based on a PI3K p.N345K mutation not identified by TGP for an invasive ductal carcinoma and a PTEN p.M1L for a pyloric adenocarcinoma.

**Figure 1:**
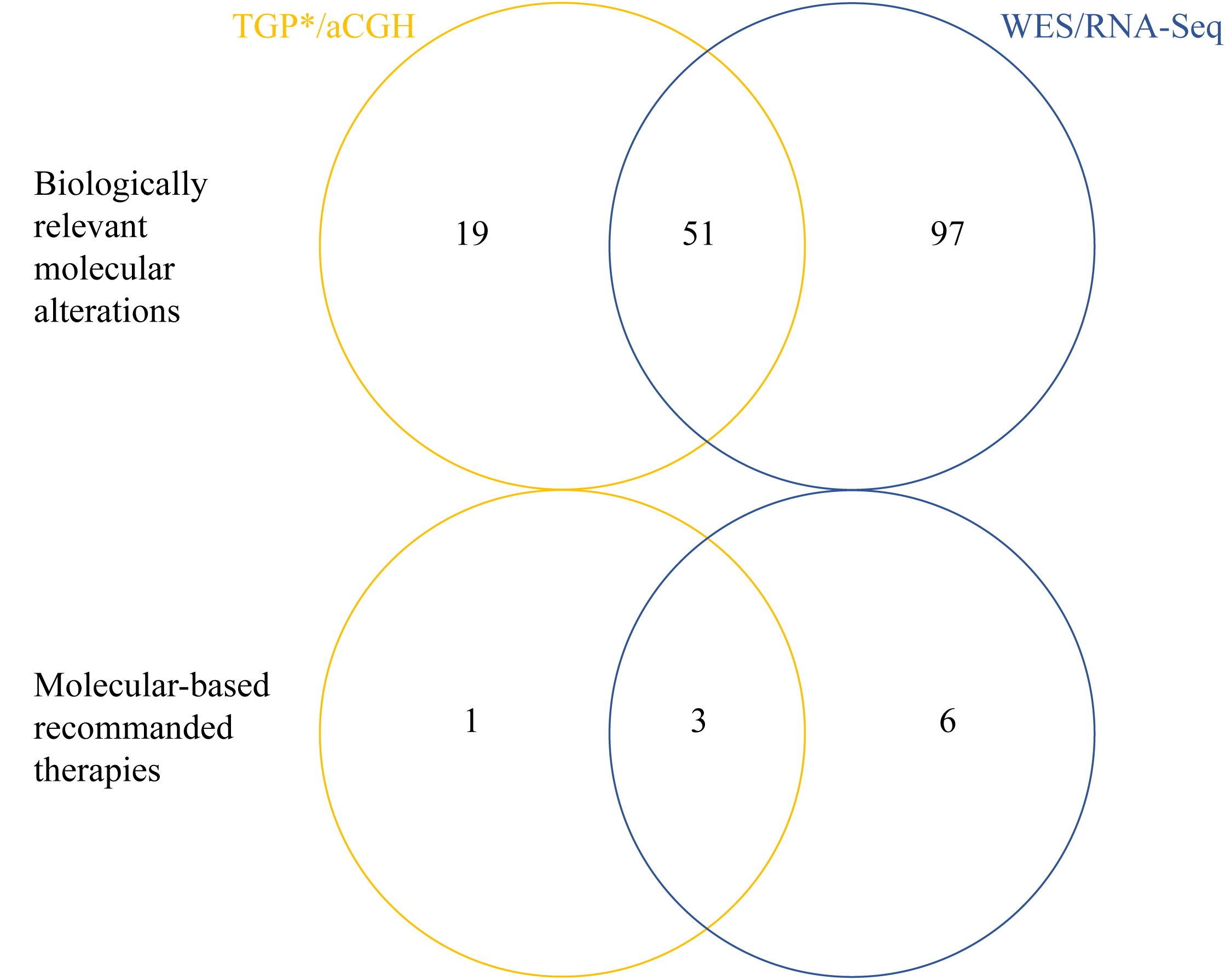
Venn diagram of biologically relevant molecular alterations identified with TGP/aCGH versus WES/RNA-Seq in advanced and refractory patients with no molecular-based recommended therapy. *TGP raw data for the 50 selected patients were reanalyzed and a new report was issued. Both the TGP/aCGH and WES/RNA-Seq reports were presented at the MTB.

## Discussion

Molecular analysis by WES/RNA-Seq is now available in routine for diagnosis and theranostic purposes to increase the rate of MBTR for patients with advanced cancer. To our knowledge, this is the first report comparing the percentage of candidate patients for a MBTR using both TGP/aCGH and WES/RNA-Seq available in all patients. As expected, WES/RNA-Seq led to the identification of more molecular alterations but most were not used for MBTR in the absence of documented clinical significance. However, a numerically higher rate of MBTR was recommended compared to TGP/aCGH. The translation of these recommendations into clinical benefit for the patients remains to be determined. The prospective randomized trial PROFILER02 trial (NCT03163732) has been completed and compares the value of a narrow-vs. larger TGP.

Only 51 (30%) molecular alterations were common to both TGP/aCGH and WES/RNA-Seq. These discrepancies may be explained by tumor heterogeneity: although the same tumor was analyzed, nucleic acids were extracted from a FFPE sample (TGP/aCGH) and from a frozen sample (WES/RNA-Seq). As expected, some molecular alterations were missed by TGP because genes were not included in the panel, or because no fusion can be studied with TGP. Discrepancies between TGP and WES/RNAseq may also be related to lower sequencing depth (false negatives), and to the subtraction of constitutional variants (true negatives).

Other groups reported an “actionable” alteration ranging from 38% to 57% of patients suitable to molecular-based therapy after extensive genomic analysis [12, 13]. However, the definition of “actionability” of a given molecular alteration remains unclear. In the study, we selected patients who were given no recommendation in the course of the trial based on TGP/aCGH, possibly explaining the low rate of patients with MBTR based on WES/RNA-Seq [9/50 (18%)].

## Conclusion

In this work, WES/RNA-Seq analysis resulted in a significantly superior but modest improvement of the number of MBTR compared to TGP/aCGH. Discrepancies were observed between the two tests, owing possibly to sample quality bias, and subclonal analysis. As more knowledge is gained on the significance of individual and combined mutations based on WES/RNA-Seq, a careful clinical evaluation of the utility of WES/RNA-Seq for the management of cancer patients with advanced and refractory disease must be undertaken to further compare the utility of narrow panels versus broader but more expensive approaches.

## Supporting information

Supplementary results

## Data Availability

All data produced in the present work are contained in the manuscript

## Acknowledgement

The authors thank the Biobank at Centre Léon Bérard (BB-0033-00050, CRB Centre Léon Bérard, Lyon, France).

## Supplementary Material

**Supplementary Figure**: Bioinformatic workflow to classify molecular alterations according to ESCAT [11]

**Supplementary Method**: Whole exome and RNA sequencing - Bioinformatics analysis

**Supplementary Table 1**: Gene list of the targeted 90-gene panel used in the PROFILER study [7]

**Supplementary Table 2**: Detailed molecular alterations identified in the 50 patients included in the study based on TGP or WES/RNA-Seq

## References

1. Drilon, A., et al., Efficacy of Larotrectinib in TRK Fusion-Positive Cancers in Adults and Children. N Engl J Med, 2018. 378(8): p. 731–739.

2. Le, D.T., et al., Mismatch repair deficiency predicts response of solid tumors to PD-1 blockade. Science, 2017. 357(6349): p. 409–413.

3. Marcus, L., et al., FDA Approval Summary: Entrectinib for the Treatment of NTRK gene Fusion Solid Tumors. Clin Cancer Res, 2021. 27(4): p. 928–932.

4. Le Tourneau, C., et al., Molecularly targeted therapy based on tumour molecular profiling versus conventional therapy for advanced cancer (SHIVA): a multicentre, open-label, proof-of-concept, randomised, controlled phase 2 trial. Lancet Oncol, 2015. 16(13): p. 1324–34.

5. Massard, C., et al., High-Throughput Genomics and Clinical Outcome in Hard-to-Treat Advanced Cancers: Results of the MOSCATO 01 Trial. Cancer Discov, 2017. 7(6): p. 586–595.

6. Stockley, T.L., et al., Molecular profiling of advanced solid tumors and patient outcomes with genotype-matched clinical trials: the Princess Margaret IMPACT/COMPACT trial. Genome Med, 2016. 8(1): p. 109.

7. Tredan, O., et al., Molecular screening program to select molecular-based recommended therapies for metastatic cancer patients: analysis from the ProfiLER trial. Ann Oncol, 2019. 30(5): p. 757–765.

8. Jardim, D.L., et al., Impact of a Biomarker-Based Strategy on Oncology Drug Development: A Meta-analysis of Clinical Trials Leading to FDA Approval. J Natl Cancer Inst, 2015. 107(11).

9. Schwaederle, M., et al., Impact of Precision Medicine in Diverse Cancers: A Meta-Analysis of Phase II Clinical Trials. J Clin Oncol, 2015. 33(32): p. 3817–25.

10. Schwaederle, M., et al., Association of Biomarker-Based Treatment Strategies With Response Rates and Progression-Free Survival in Refractory Malignant Neoplasms: A Meta-analysis. JAMA Oncol, 2016. 2(11): p. 1452–1459.

11. Mateo, J., et al., A framework to rank genomic alterations as targets for cancer precision medicine: the ESMO Scale for Clinical Actionability of molecular Targets (ESCAT). Ann Oncol, 2018. 29(9): p. 1895–1902.

12. Beg, S., et al., Integration of whole-exome and anchored PCR-based next generation sequencing significantly increases detection of actionable alterations in precision oncology. Transl Oncol, 2021. 14(1): p. 100944.

13. Koeppel, F., et al., Standardisation of pathogenicity classification for somatic alterations in solid tumours and haematologic malignancies. Eur J Cancer, 2021. 159: p. 1–15.

