## Supplementary methods for "Added-value of whole exome and RNA Sequencing in advanced and refractory cancer patients with no molecular-based treatment recommendation based on a 90-gene panel"

**Supplementary Method**: Whole exome and RNA sequencing - Bioinformatics analysis

**Whole exome and RNA sequencing**

Whole exome was captured from frozen tumor sample and paired constitutional DNA and libraries were prepared using Agilent SureSelect XT low input V6 + UTR (91 Mb) (Agilent Technologies, Santa Clara, CA, USA). Libraries were sequenced on a on a NovaSeq6000 Illumina sequencer in 100 bp paired-end (Illumina, Santa Diego, CA, USA). Bioinformatics processing is detailed in Supplementary Methods.

For RNA sequencing, libraries were prepared with TruSeq Stranded mRNA kit following recommendations (Illumina, Santa Diego, CA, USA. and sequenced on a NovaSeq6000 sequencer in 75 bp paired-end.

**Whole exome sequencing bioinformatics analysis**

Whole exome was captured from 200 ng of tumor from frozen tumor and paired constitutional DNA and libraries were prepared using Agilent SureSelect XT low input V6 + UTR (68 Mb). Libraries were sequenced of subsequent libraries was performed using the NovaSeq6000 Illumina sequencer in 100 bp paired-end. Bioinformatics processing is detailed in Supplementary Methods.

All exome sequencing data were mapped to the *hg38* human reference genome with *bwa aligner v0.7.15-r1140* (<https://github.com/lh3/bwa>). Alignments were sorted and duplicates were marked using *biobambam v2.0.79* (<https://gitlab.com/german.tischler/biobambam2>).

Somatic point mutations and short indels were called using *Mutect2* module from *GATK v4.1.0.0-37-g80ab76b* (<https://github.com/broadinstitute/gatk>). The genome aggregation database *gnomAD v2.1.1* was used as a resource to calibrate variant calling model in Mutect2. Also, a panel of 50 normal samples sequenced with the same capture kit and on the same sequencing machine (NovaSeq6000) was used to remove recurring sequencing artifacts. All variants were annotated using the ensembl-vep v90 annotation tool which integrates *COSMIC v81* and *dbSNP v150* resources in its cache.

Somatic copy number alterations were called using *facets v0.5.14* which also provides estimations of the tumor purity and ploidy. Only amplifications and regions of homozygous deletions were considered to be evaluated in this study. A gene was claimed amplified (AMP) if it has at least 8 copies in tumor cells. A gene was claimed deleted (HDEL) if it has no remaining copy in tumor cells.

Tumor mutational burden (TMB) was computed from non-synonymous variants (point mutations or indels) on the exome target with at least a variant allele frequency (vaf) of 10% and is reported in mutation per megabase (mut/Mb).

**RNA sequencing bioinformatics analysis**

Alignments were performed using STAR on the GRCh38 version of the human reference genome. Number of duplicate reads were assessed using PICARD tools. Fusion transcripts were called by five different algorithms, including STAR-Fusion, FusionMap, FusionCatcher, TopHat-Fusion, and EricScript.

**Gene panel and aCGH bioinformatics analysis**

All tumor samples were mapped to the hg37 human reference genome with *bwa v0.7.12*. Alignments were sorted and duplicates were marked using *Picard tools v1.121* (<https://broadinstitute.github.io/picard/>) . Re-alignment around known indels and base quality score recalibration were then performed with *GATK v3.5-0*.

Somatic point mutations and short indels were called using *UnifiedGenotyper* module of *GATK v3.5-0*. All identified variants were then annotated with *ANNOVAR v02_2016* which included *dbSNP v135* and *COSMIC v64* resources. The variants were then filtered according to their frequency (>5% for point mutations and >10% for indels), their local coverage (>50X for point mutations and >100X for indels) and their effect (non-synonymous, splicing, not polymorphism, known hotspot).

aCGH analyses were carried out using the *Agilent Genomic Workbench software v7.0*. The identification of aberrant copy number segments was based on ADM-2 segmentation algorithm with a threshold of 7. A null log2 ratio corresponds to a balanced tumor/normal ratio. By default, for samples with a high percentage of tumor cells (≥80%), low-level and high-level copy number gains / losses were defined as a |log2 (ratio)| >0.25 and 1.5, respectively. Gene Amplification: log2 ratio ≥2. These thresholds were adjusted for samples with a lower percentage of tumor cells. When needed, aCGH results were confirmed by FISH analysis.

**Inferring clinical actionability of variants**

All variants found either in WES or panel sequencing data and gene fusions found in RNAseq data were submitted anonymously to the *Cancer Genome Interpreter* (CGI) web interface. This resource is a clinical knowledge database manually curated and maintained by clinical and scientific experts that allows to map gene alterations to levels of evidence supporting responsiveness or resistance of a given treatment in a given pathology.

CGI website: <https://www.cancergenomeinterpreter.org>

Cancer Biomarkers database: <https://www.cancergenomeinterpreter.org/biomarkers>
